# Impact of a regional educational advertising campaign on harm perceptions of e-cigarettes, prevalence of e-cigarette use, and quit attempts among smokers

**DOI:** 10.1101/19001610

**Authors:** Harry Tattan-Birch, Sarah E Jackson, Charlotte Ide, Linda Bauld, Lion Shahab

## Abstract

**Background:** We evaluated how effective an advertising campaign that was piloted by Cancer Research UK in January/February 2018 was at promoting quit attempts by increasing awareness of the relative harms of e-cigarettes compared with smoking.

**Methods:** Adults (≥16 years, *n* = 2217) living in Greater Manchester (campaign region) and Yorkshire & Humber and the North East of England (control regions) completed cross-sectional surveys immediately before and after the campaign period. Surveys measured socio-demographics, perceptions and use of e-cigarettes, and motivation and attempts to quit smoking. We tested interactions between time (pre, post) and region (campaign, control).

**Results:** 36.7% (95% CI 33.0% – 40.6%) of those in the intervention region recognised the campaign. In the general population, interactions were non-significant for all outcomes except for perception of e-cigarettes as effective cessation aids, with smaller increases from pre-to post-campaign in the campaign (49.9% to 54.0%) compared with the control region (40.5% to 55.0%; OR = 0.66, 95% CI 0.45 – 0.98). Among smokers, motivation to quit increased in the intervention region (44.0% to 48.0%) but decreased in the control region (40.5% to 21.5%; OR = 2.97, 95% CI 1.25 – 7.16), with no other significant differences between regions over time. A Bayesian analysis confirmed that non-significant results were inconclusive.

**Conclusions:** Compared with the control region, the campaign was associated with an increase in smokers’ motivation to quit but a smaller increase in adults’ perception of e-cigarettes as an effective cessation aid. There was insufficient evidence to determine whether the campaign affected other outcomes.

## INTRODUCTION

Tobacco smoking is associated with substantial risk of premature morbidity and mortality (1) and is the leading preventable cause of cancer, causing around 7 in 10 lung cancer cases in the UK (2). This excess risk is primarily driven by the inhalation of toxins contained within the smoke (3) and could be mitigated by quitting or switching to nicotine products that do not contain tobacco or require combustion (4). Electronic cigarettes (e-cigarettes) are handheld electronic devices that allow the user to inhale nicotine in a vapour, without tobacco or combustion, thereby reducing nicotine cravings and withdrawal symptoms (5). However, public perceptions of e-cigarettes are inaccurate and have worsened over recent years even among smokers (5,6), which may discourage use. Increasing awareness of the relative harms of e-cigarettes and tobacco smoking is important to enable smokers to make informed decisions about product use. In this study, we evaluate the effectiveness of an advertising campaign designed to address misperceptions around e-cigarette harms relative to tobacco smoking.

Following a period of rapid growth in the prevalence of e-cigarette use (‘vaping’) from 2011 to 2014, e-cigarettes are now used by around 2.6 million people in England (20% of smokers) and are the most common aid to smoking cessation, used in over one third of quit attempts (7). Evidence from several randomised controlled trials indicates that using e-cigarettes in a quit attempt increases the chances of successful cessation (8,9). On a population level, the rise in use of e-cigarettes in England and the United States has been associated with increases in the overall success rate of quit attempts in the population (10,11), contributing to continued declines in smoking prevalence (12). While the prevalence of e-cigarette use in England has remained fairly stable since late 2013, use among long-term ex-smokers has continued to rise and some data suggest that there are now more ex-smokers than smokers using e-cigarettes (14).

Although the long-term effects of using e-cigarettes are unclear as they are a relatively new product, a growing body of evidence demonstrates that they are much less harmful than smoking tobacco (5,15). Toxicology testing has shown that while e-cigarettes can be used to obtain similar levels of nicotine to combustible tobacco, switching to e-cigarettes can significantly reduce levels of measured carcinogens and toxins relative to smoking, with differences observed within a matter of weeks (16–18). Moreover, lower levels of carcinogens and toxins have been observed among long-term e-cigarette users (≥6 months) compared with current cigarette smokers (15).

However, surveys have indicated that public perceptions of the relative harms of using e-cigarettes compared with smoking tobacco are generally inaccurate. In 2017, less than half (44%) of adults in Great Britain believed that e-cigarettes are less harmful than smoking, the lowest percentage since monitoring started in 2013 (5,6). Just 13% correctly identified e-cigarettes as being a lot less harmful than smoking (5,6). The majority of adults perceived e-cigarettes to be equally harmful (23%), more harmful (3%), or were unsure about the relative risk (29%), with the percentage who believed they were more harmful the highest on record (5,6). Inaccurate perceptions of harm may undermine the potential public health benefits of e-cigarettes as an option for smoking cessation. If smokers are unaware that e-cigarettes offer a less harmful alternative, they may be less likely to try using e-cigarettes to quit. Thus, there is a need for effective interventions to increase the accuracy of perceptions of the relative harm associated with e-cigarettes, particularly among smokers from groups with high prevalence of smoking or low likelihood of quitting.

In addition to nationwide efforts to correct these misperceptions, Cancer Research UK, the world’s largest cancer charity, developed an advertising campaign that aimed to increase awareness of the relative harms of e-cigarettes compared with tobacco. The campaign was targeted at smokers aged 25-55 years, with a focus on those who wanted to stop and had never tried e-cigarettes. Lower socio-economic groups were also targeted because people with greater disadvantage are more likely to smoke and less likely to use e-cigarettes than those who are more advantaged (19) (although recent data suggest this disparity is narrowing (20)). The campaign was piloted in Greater Manchester over a four-week period in early 2018, with surveys conducted before and after the campaign period to evaluate changes in perceptions and use of e-cigarettes and quit attempts. For evaluation purposes, surveys were also conducted in the designated control regions Yorkshire & Humber and the North East. These were chosen as comparison regions because they share a similar demographic profile to Greater Manchester in terms of smoking prevalence, deprivation, and age.

This study used data from these surveys to evaluate the extent to which the campaign was effective in:

1. Reaching members of the target group, C2DE smokers aged 25-55 years,
2. Increasing awareness of the harms of e-cigarettes relative to smoking tobacco,
3. Improving smokers’ attitudes to using e-cigarettes,
4. Promoting actual use of e-cigarettes by smokers, and
5. Increasing motivation and attempts to quit among smokers.

## METHODS

### Design and setting

Cross-sectional surveys were conducted on samples of adults (≥16 years) living in Greater Manchester (intervention region) and Yorkshire & Humber and the North East (control region) in two waves, one before (11 December 2017 – 4 January 2018) and one after (22 February – 13 March 2018) the intervention period. All research by Cancer Research UK is carried out according to the Market Research Society Code of Conduct. This study used anonymized records and datasets available from Cancer Research UK who had already acquired appropriate permissions from participants.

### Registration

The preregistered analysis plan, data and code used to generate results is available on the Open Science Framework (https://osf.io/uswpj/).

### Participants

Participants (n=2,217) were recruited via online consumer panels and incentivised (with points that were accrued across multiple surveys and could be exchanged for cash or online vouchers) to complete a roughly 15-minute online survey. Quota sampling was used to match the four survey samples (intervention and control, pre- and post-intervention) on age group (25% 16-29, 25% 30-44, 25% 45-60, 25% >60 years), sex (50% male, 50% female), social grade (based on the occupational group of the household’s chief income earner; 50% ABC1, 50% C2DE), and smoking status (19% current smoker, 25% ex-smoker, 56% non-smoker; based on local prevalence data (21)). A boost sample (n=580) of C2DE smokers in the Greater Manchester region was recruited to facilitate more detailed examination of this target group.

### Intervention

The intervention was an advertising campaign developed by Cancer Research UK that aimed to correct misperceptions about relative harms of using e-cigarettes compared with smoking tobacco. The main messages of the campaign were:

i. Research so far shows that vaping is far less harmful than smoking.
ii. E-cigarettes don’t contain tobacco, which is proven to cause cancer. They do contain nicotine, which is addictive, but isn’t responsible for the major health harms from smoking.
iii. Many people are now using e-cigarettes to help them stop smoking.

The campaign’s activities included:

i. An outdoor advertising campaign, including adverts on buses, billboards, bus stops, phone kiosks and washroom posters across Greater Manchester;
ii. Regional press coverage in Greater Manchester; and
iii. Facebook adverts run UK-wide (excluding control regions Yorkshire & Humber and the North East).

The campaign was piloted in Greater Manchester over a four-week period between 15 January and 18 February 2018. Greater Manchester was selected for the pilot study as a region with particularly high smoking prevalence and deprivation levels (22,23), and a commitment (within the Greater Manchester Tobacco Control Plan ‘Making Smoking History’) to develop innovative e-cigarette friendly policies, services, and offers (24).

### Measures

The exposure variable was region, with Greater Manchester (campaign region) coded 1 and Yorkshire & Humber and the North East (control region) coded 0.

A number of relevant outcome variables were evaluated. Recognition of the campaign materials was assessed among all respondents in Greater Manchester (including the boost sample) at follow-up by presenting them with images of the campaign poster, out of home material, social media infographics, and PR content and asking for each of the four types of content: “Before today, had you seen this content?” Those who respond yes to any of the four questions (i.e. reported having seen any of the content) were coded 1 and those who respond no to all questions (i.e. reported not having seen any of the content) were coded 0.

Harm perceptions of e-cigarettes were assessed among all respondents with the question: “Which of the following best describes what you think about e-cigarettes? Responses were dichotomised to distinguish between those who perceive e-cigarettes as less harmful or a lot less harmful than cigarettes and all other response options.

Perception of e-cigarettes as an effective aid to cessation was assessed among all respondents with the question: “To what extent do you think e-cigarettes are an effective aid to stop smoking regular cigarettes?”. Responses were recorded on a 6-point Likert-scale (strongly disagree to strongly agree). These were dichotomised to distinguish between those who agree that e-cigarettes are an effective aid to cessation and all other response options.

Whether respondents would consider using e-cigarettes as a cessation aid was assessed among current smokers with the question: “To what extent are you likely to consider using e-cigarettes as an aid to stop smoking regular cigarettes in the future?”. Responses were recorded on a 6-point Likert-scale (very unlikely to very likely). These were dichotomised to distinguish between those who would be likely to use e-cigarettes as an aid to cessation and all other response options.

Whether respondents would recommend e-cigarettes as a cessation aid to someone else was assessed among all respondents with the question: “To what extent are you likely to recommend e-cigarettes to someone you know as an aid to stop smoking regular cigarettes?” Responses were recorded on a 6-point Likert-scale ranging (very unlikely to very likely). These were dichotomised to distinguish between those who would be likely to recommend e-cigarettes as an aid to cessation and all other response options.

E-cigarette use was assessed among current and ex-smokers with the question: “Have you tried an electronic cigarette or vaping device in the last two months?” with yes coded 1 and no coded 0. Frequency of current e-cigarette use was assessed among those who report having used an e-cigarette in the last two months with the question: “How often, if at all, do you currently use an electronic cigarette or vaping device?” Responses were dichotomised to distinguish between those who use e-cigarettes daily and all other response options.

Motivation to stop smoking was assessed among current smokers using the Motivation to Stop Scale (25), Responses were dichotomised to distinguish between those with high motivation (i.e. intend to stop within the next 3 months) and all other response options (26,27).

Quit attempts were assessed among current and ex-smokers with the question: “Have you been attempting to quit smoking in the last 4 to 8 weeks? Please include any attempts you’re currently making” with yes coded 1 and no coded 0.

Covariates included: age group (16-29, 30-44, 45-60, >60 years), sex (male, female), and social grade (ABC1, C2DE). Smoking status was measured by asking “Which of the following BEST applies to you?”. Those who responded “I smoke, but not everyday” or “I smoke everyday” were considered current smokers and all others were considered non-smokers.

### Analyses

We used descriptive statistics to summarise responses to the outcome variables in the intervention and control regions before and after the intervention. Differences in recognition of the campaign materials at follow-up between target (i.e. age 25-55 years and social grades C2DE) and non-target groups (i.e. age <25 or >55 and social grades ABC1) were analysed using logistic regression. This analysis included participants from the boost sample. Interactions between time (pre-intervention, post-intervention) and region (campaign, control) for all outcomes of interest were analysed using generalised estimating equations. Participants recruited as part of the boost sample were excluded to maintain comparability of the intervention and control samples. Among all respondents, we tested whether there was a greater change in the intervention region versus the control region from pre-to post-intervention in perceptions of e-cigarettes. We also tested the change in perceptions of e-cigarettes among a sample of current smokers, alongside other relevant outcomes such as motivation to quit, e-cigarette use, and quit attempts. Among current smokers who reported using e-cigarettes at all in the past two months, we tested whether there was a greater change in daily e-cigarette use. All analyses controlled for age, sex, and social grade.

We calculated Bayes factors (BF) for non-significant interactions to determine whether they provide evidence for no effect (BF < 1/3) when compared to the alternative hypothesis or indicate data insensitivity (BF≥1/3 and <3). The alternative hypothesis was modelled as a half-normal distribution centred on zero, with a standard deviation equal to the expected effect size. The expected effect size (odds ratio (OR) = 1.24) was used based on previous literature (28,29). Sensitivity analyses were performed using smaller (OR = 1.10) and larger (OR = 1.40) expected effect sizes to test how robust conclusions were to changes in priors.

## RESULTS

Socio-demographic characteristics of the samples in each region pre- and post-campaign are shown in Table 1. Over a third (36.7%, 95% CI 33.0% – 40.6%) of participants in the post-campaign Manchester sample recognised at least one of the campaign materials. There was no significant difference in recognition between target participants (C2DE smokers aged 25-55 years) and others (38.3% vs. 35.8%, OR= 1.17, 95% CI 0.83 – 1.65). The aspect of the campaign with the largest reach was the bus stop poster with 28.7% of participants reporting recognising it.

**Table 1.** Sample Characteristics

|  | Control Regions |  | Campaign Region |  |
| --- | --- | --- | --- | --- |
|  | Before, % (n) | After, % (n) | Before, % (n) | After, % (n) |
| Smoking status |  |  |  |  |
| Non-smoker | 73.6 (309) | 80.4 (325) | 63.0 (260) | 81.3 (325) |
| Smoker | 26.4 (111) | 19.6 (79) | 37.0 (153) | 18.8 (75) |
| Social Grade |  |  |  |  |
| ABC1 | 43.6 (183) | 50.2 (203) | 42.6 (176) | 50.8 (203) |
| C2DE | 56.4 (237) | 49.8 (201) | 57.4 (237) | 49.3 (197) |
| Age (years) |  |  |  |  |
| 16-29 | 18.3 (77) | 23.5 (95) | 21.1 (87) | 24.5 (98) |
| 30-44 | 28.8 (121) | 26.5 (107) | 31.5 (130) | 26.3 (105) |
| 45-60 | 35.2 (148) | 24.5 (99) | 33.4 (138) | 25.3 (101) |
| >60 | 17.6 (74) | 25.5 (103) | 14.0 (58) | 24.0 (96) |

Table 2 shows changes in outcomes over time by region. In the general population, interactions between time and region were non-significant for all outcomes except for perception of e-cigarettes as effective cessation aids, with smaller increases from pre- to post-campaign in the campaign region (49.9% to 54.0%) compared with the control region (40.5% to 55.0%; OR = 0.66, 95% CI 0.45 – 0.98, p = .04). In current smokers, the only significant time by region interaction was for motivation to quit, which increased in the intervention region (44.0% to 48.0%) but decreased in the control region (40.5% to 21.5%; OR = 2.97, 95% CI 1.25 – 7.16, p = .01).

**Table 2:** Results of generalised estimating equations testing interactions between region (campaign, control) and time (pre- and post-campaign) for e-cigarette perceptions and use, motivation to quit, and incidence of quit attempts.

|  | **Control Region, % (n)** | **Campaign Region, % (n)** | **Main Effect Region, OR (95% CI)** | **Main Effect Time, OR (95% CI)** | **Interaction Group and Time, OR (95% CI)** |
| --- | --- | --- | --- | --- | --- |
| **Outcomes among all Participants** |  |  |  |  |  |
| Perceptions of e-cigarettes as less harmful than conventional cigarettes |  |  |  |  |  |
| Pre | 48.3 (203) | 55.0 (227) |  |  |  |
| Post | 57.5 (234) | 57.8 (231) | 1.29 (0.98 - 1.69), $p = .07$ | 1.46 (1.10 - 1.92), $p < .01$ | 0.76 (0.51 - 1.13), $p = .18$, BF = 0.36 |
| Perception of e-cigarettes as effective cessation aid |  |  |  |  |  |
| Pre | 40.5 (170) | 49.9 (206) |  |  |  |
| Post | 55.0 (222) | 54.0 (216) | 1.44 (1.10 - 1.90), $p < .01$ | 1.82 (1.38 - 2.41), $p < .001$ | 0.66 (0.45 - 0.98), $p = .04$ |
| Likely to recommend e-cigarettes as cessation aid |  |  |  |  |  |
| Pre | 28.6 (120) | 34.4 (142) |  |  |  |
| Post | 29.2 (118) | 36.0 (144) | 1.26 (0.94 - 1.70), $p = .13$ | 1.04 (0.76 - 1.41), $p = .82$ | 1.07 (0.70 - 1.64), $p = .74$, BF = 0.86 |
| **Outcomes among Smokers** |  |  |  |  |  |
| Perceptions of e-cigarettes as less harmful than conventional |  |  |  |  |  |
| cigarettes |  |  |  |  |  |
| Pre | 43.2 (48) | 32.9 (26) |  |  |  |
| Post | 56.8 (63) | 67.0 (53) | 0.91 (0.55 - 1.50), p = .71 | 3.00 (1.24 - 7.60), p = .02 | 0.97 (0.41 - 2.3), p = .95, BF = 0.88 |
| Perception of e-cigarettes as effective cessation aid |  |  |  |  |  |
| Pre | 51.4 (57) | 54.2 (83) |  |  |  |
| Post | 55.7 (44) | 64.0 (48) | 1.08 (0.65 - 1.78), p = .76 | 0.65 (0.23 - 1.79), p = .40 | 1.36 (0.59 - 3.17), p = .47, BF = 1.18 |
| Likely to use e-cigarette as cessation aid |  |  |  |  |  |
| Pre | 42.3 (47) | 49.7 (76) |  |  |  |
| Post | 49.4 (39) | 56.0 (42) | 1.32 (0.80 - 2.2), p = .28 | 0.98 (0.35 - 2.72), p = .97 | 0.96 (0.42 - 2.20), p = .92, BF = 0.86 |
| Likely to recommend e-cigarettes as cessation aid |  |  |  |  |  |
| Pre | 38.7 (43) | 43.8 (67) |  |  |  |
| Post | 40.5 (32) | 56.0 (42) | 1.2 (0.72 - 2.01), p = .47 | 1.09 (0.59 - 2.02), p = .78 | 1.44 (0.63 - 3.32), p = .39, BF = 1.25 |
| High motivation to stop smoking |  |  |  |  |  |
| Pre | 40.5 (45) | 44.0 (67) |  |  |  |
| Post | 21.5 (17) | 48.0 (36) | 1.11 (0.67 - 1.84), p = .68 | 0.38 (0.19 - 0.74), p < .01 | 2.97 (1.25 - 7.16), p = .01 |
| Used E-cigarette in past 2 months |  |  |  |  |  |
| Pre | 41.5 (44) | 44.1 (63) |  |  |  |
| Post | 42.5 (31) | 56.7 (38) | 1.12 (0.67 - 1.88), p = .67 | 1.10 (0.59 - 2.06), p = .76 | 1.51 (0.64 - 3.60), p = .35, BF = 1.29 |
| Make a quit attempt in past 2 months |  |  |  |  |  |
| Pre | 29.7 (33) | 41.8 (64) |  |  |  |
| Post | 30.4 (24) | 54.7 (41) | 1.67 (0.99 - 2.85), p = .06 | 0.97 (0.50 - 1.85), p = .93 | 1.60 (0.68 - 3.79), p = .28, BF = 1.38 |
| Use e-cigarettes daily |  |  |  |  |  |
| Pre | 13.0 (6) | 28.9 (22) |  |  |  |
| Post | 28.2 (11) | 35.7 (15) | 2.37 (0.97 - 6.20), p = .07 | 1.55 (0.50 - 4.79), p = .44 | 0.78 (0.19 - 3.17), p = .72, BF = 0.88 |

Notes: *OR* odds ratio, *CI* confidence interval, *BF* Bayes factor

Bayes factors were calculated for non-significant results to determine whether the data were insensitive or provided evidence for no effect. All Bayes factors were between a third and three, suggesting that there was insufficient evidence to determine the impact of the campaign across these outcomes. Sensitivity analyses that used both higher and lower expected effect sizes provided the same conclusion for all but one of the interactions (see supplementary material), suggesting that there was insufficient evidence to reject or accept either the null or alternative hypotheses.

## DISCUSSION

An awareness campaign to improve harm perceptions of e-cigarettes relative to smoking, developed by Cancer Research UK, was recognised by over a third of participants in a post-campaign evaluation. Recognition was not significantly higher in the target group of low socio-economic status (C2DE) smokers aged 25-55 years. When compared with smokers from a control region, smokers who were exposed to the campaign reported a more positive change in motivation to quit from before to after the campaign was run. However, among all participants, perceptions of e-cigarettes as an effective cessation aid improved more in the control than campaign region. There was insufficient evidence to determine the impact of the campaign on other outcomes, including the campaign’s primary objective: harm perceptions of e-cigarettes relative to smoking.

Past mass media campaigns have successfully altered the public’s attitudes towards smoking, improved motivation to quit smoking, and reduced smoking prevalence (30). This campaign also aimed to promote quitting through the most common cessation aid in the UK: e-cigarettes (7). Given that the campaign only ran for one month, reach was impressive with more than one in three of those surveyed recognising at least one of the advertisements. This is comparable to the level of recognition recorded from other smoking cessation campaigns than ran for a much longer duration (31,32). The similarly high level of recognition found among C2DE smokers aged 25-55 years and other populations suggests that, while the campaign was effective at reaching this target group, it was similarly successful at reaching other groups.

Motivation to quit smoking tends to be high at the start of a new year and subsequently drops off throughout January (33). We found that those in the campaign region were insulated from this drop. The campaign’s promotion of e-cigarettes as a cessation aid, which are widely available and often quicker to access than other cessation aids such as pharmacotherapy on prescription, may have contributed to this. Motivation to quit is a strong predictor of future quit attempts (34). Therefore, if the difference in motivation between regions was caused by the campaign, there is a rationale to expand it to other regions. However, other unmeasured influences may also have contributed towards these different trends across regions.

Perceptions of e-cigarettes as effective cessation aids increased in both regions from pre-to post-campaign, but showed a significantly greater increase in the control region. At baseline, perceptions of e-cigarettes as effective cessation aids were much higher in the campaign region. Therefore, the smaller increase in this region may represent a ceiling effect, where the high initial level of this variable meant that it was unresponsive to interventions aimed to raise it. Conversely, low baseline effectiveness perceptions in the control region meant other campaigns aimed at correcting these misperceptions — such as one run by Public Health England (35) over the same time period — would likely have had a greater impact, resulting in similar levels of perceptions at follow-up.

Bayes factors indicated data were insensitive to detect significant associations for most outcomes. Effects of mass media advertising campaigns tend to be small, especially if they are run for a short period of time (29). Therefore, the lack of evidence to determine whether changes in the other outcomes differed between regions was likely due to a lack of statistical power. We may have been able to detect the campaign’s impact on these outcomes if it ran for a longer time period, or if the sample size was larger.

A strength of this study was that we were able to adjust for trends in outcomes over time by using a comparison region containing populations who were not exposed to the campaign. A pre-post analysis solely in the campaign region would have been unable to determine what effects were caused by seasonal changes, which are especially variable from December to February (33). Nonetheless, the study design used had several limitations. Most significantly, we had to assume that the trends in outcomes would be identical across groups were it not for the influence of the advertising campaign. For this to be true, any unmeasured variables that influence outcomes must either be constant over time or equal across groups, which is unlikely to be the case. For instance, over the campaign period, there were other interventions in place (in addition to the Cancer Research UK campaign) that aimed to alter public perceptions of e-cigarettes (35). These may have had a greater impact on outcomes in the control region due to lower baseline perceptions. Moreover, there is regular news coverage both supporting and opposing e-cigarette use, which could also have differentially affected trends across regions. To better account for these unobserved variables, a longer time-series analysis or cluster randomised-controlled trial would be required. Another limitation is that all data were self-reported, which introduces scope for error and bias. However, due to the low demand characteristics in the study, this is unlikely to significantly influence our results. Issues also arise from the sample used. Firstly, unlike a true pre-post study, different individuals were recruited in baseline and follow-up samples. This introduces bias as these samples may vary on important characteristics. Secondly, sample size was insufficient to detect even relatively large changes in outcomes. Future research, with a larger sample and more robust design, would be useful to determine whether similar advertising campaigns can impact harm perceptions towards e-cigarettes when compared to smoking.

## CONCLUSIONS

A regional educational advertising campaign in England was associated with increased motivation to quit among smokers exposed to the campaign, when compared with those in an unexposed region. It was also associated with a smaller increase in perceptions of e-cigarettes as an effective cessation aid among the general population, which may be due to different baseline perceptions across regions. A Bayesian analysis showed that the impact of the campaign on the other outcomes was inconclusive, which may indicate that the campaign needed to run across a longer period to cause a detectable effect.

## Acknowledgements

This pilot campaign was funded by Cancer Research UK. CI designed and managed the evaluation of the campaign. SJ drafted the introduction and methods sections of this manuscript, while HTB ran the data analysis and produced the results and discussion. LS, CI and LB provided amendments to the manuscript. All authors read and approved the final version.

